# Quantitative deep motor phenotyping of 102 people with *SCN2A, KCNQ2, CSNK2A1, CACNA1A* or *SCN8A*-related disorders

**DOI:** 10.64898/2026.11.03.26365785

**Authors:** Sunanjay Bajaj, Tobias Brünger, Mariana Fauteux, Roshirl Fracisco, Manuela Ochoa-Urrea, Chandler Sargent, Alison Merket, Anu Cherukara, Emile Moura, Althea Smith, Ludovica Montanucci, Suyeon Kim, Costin Leu, Christiane Hsu, Gary Taylor, Vanessa Kasto, Walid Fazeli, Jaehyung Lim, M. Scott Perry, Sarah M.L. Wilson, Sara E. McMahan, Shelley S. Varnado, Samden D. Lhatoo, Gretchen Von Allmen, Leah Schust Myers, Shawn Egan, Brooke Babineau, Scotty Sims, Kacie Craig, Lisa Manaster, Shira Johnson, Pangkong Fox, Jennifer Sills, Gabrielle Rushing, Ingrid E. Scheffer, Nivedita Thakur, Dennis Lal

**Author notes:** Equal contribution (Sunanjay Bajaj, Tobias Brünger).

## Abstract

**Background and Objectives:** Motor dysfunction is a major contributor to disability in pediatric-onset monogenic neurodevelopmental disorders (NDDs), yet systematic quantifications of motor phenotypes across development using validated clinical scales remain underdeveloped. Existing studies have classified motor phenotypes rather than quantified domain-specific severity, limiting their use as outcome measures in clinical trials. Here, we systematically quantify motor dysfunction using established clinical rating scales to explore their utility as motor endpoints.

**Methods:** In this cross-sectional observational study, 102 individuals with NDDs (0.5-38 years) secondary to pathogenic or likely pathogenic variants in five genes (*SCN2A, KCNQ2, CSNK2A1, CACNA1A, SCN8A*) and 31 unaffected sibling controls underwent standardized motor assessments using the Scale for Assessment and Rating of Ataxia, Burke-Fahn-Marsden Dystonia Scale, and the Modified Ashworth Scale. Regional and functional categorization of parameters enabled differentiated interpretation. Gene-specific patterns were evaluated using z-score standardization across the pooled affected cohort, with gene-level averaging and nonparametric bootstrap-derived 95% confidence intervals.

**Results:** Dysfunction was present in individuals with NDDs across speech (90%), coordination (87%), dystonia (83.3%), gross motor (82.4%) and spasticity (42.2%), with minimal impairment in controls (0-12.9%). We observed substantial heterogeneity across motor domains and subdomains; ataxic dysarthria had the highest score distribution (median 0.83 [IQR, 0.17-1.0]), whereas spasticity showed near-floor scores. Individuals with *SCN2A* variants demonstrated higher mean z-scores across multiple subdomains than other genes, with more consistent deviations observed in coordination and speech domains (Coordination: mean z = 0.29, 95% CI 0.03-0.54; Speech: mean z = 0.3, 95% CI 0.03-0.57), whereas domain-specific or less pronounced patterns were observed in other genes. The ataxia and dystonia scales demonstrated low floor and ceiling effects.

**Discussion:** Using three clinical scales across a broad developmental and functional range, our study moves motor phenotyping in monogenic NDDs from categorical description toward quantitative, domain-resolved severity assessment. Exploratory analysis indicates marked heterogeneity in severity profiles across genes, most notably in *SCN2A*, suggesting potential selective motor vulnerability. These findings provide evidence for the feasibility of motor phenotyping and provide preliminary, cross-sectional benchmark severity distributions. Subsequent longitudinal validation and validation in larger, longitudinal, and variant-stratified cohorts will enable natural history studies and future therapeutic trials alongside seizure outcomes.

## Introduction

Monogenic neurodevelopmental disorders (NDDs) are characterized by diverse neurological, cognitive and autonomic impairments.^1,2^ Motor dysfunction, ranging from abnormalities of tone to severe movement disorders, is a commonly observed yet poorly studied feature, despite its substantial impact on long-term functional outcomes and quality of life.^3–5^ Recent evidence suggests motor dysfunction in individuals with NDDs is diverse^6^, present across a broad age range,^7,8^ and associated with most NDD-related genes.^9,10^ Similarly, large repositories of parent-reported data such as Simons Searchlight^11^ and meta-analyses^12,13^ provide evidence for the near-universal presence of motor dysfunction in monogenic NDDs, emphasizing that it is not merely a gene- or syndrome-specific feature. It is thus imperative to develop tools to reliably detect motor dysfunction and monitor its evolution.

However, the motor outcome measures validated for NDDs remain underdeveloped, especially in contrast to seizure-related outcomes, due to several limitations of our current knowledgebase. Firstly, motor dysfunction can be subtle in severity and difficult to distinguish from seizures, complicating detection and clinical management.^14^ As such, much of the prior work has focused on identification and subtype classification, rather than on quantifying severity.^6^ The limited previous application of standardized motor rating scales across a wide age range has further restricted the quantification of motor dysfunction and inclusion of motor outcome measures in clinical trials.^15,16^ Second, monogenic NDDs are rare, constraining prior studies by cohort size. Therefore, many analyses rely on parent-reported outcomes and retrospective chart reviews.^17,18^ Due to the dependence on multi-gene cohorts with often fewer than 10 individuals per gene^6^ or larger parent-reported registries^9,11^, gene-specific patterns of motor dysfunction are not well understood either. Finally, although video-based rating approaches have been developed in rare disease contexts, these have emphasized screening and rapid identification of motor dysfunction rather than deep, multidomain severity profiling intended for endpoint selection.^19^ Ultimately, studies using uniform, validated motor assessment instruments across individuals with NDDs remain elusive.

To define severity estimates for motor dysfunction in individuals with monogenic NDDs, we quantified motor phenotypes in a large, cross-sectional cohort spanning five monogenic NDDs from infancy through adulthood using established rating scales. We provide domain-level quantification of motor dysfunction and establish genotype-associated patterns. By anchoring motor phenotyping to three clinician-assessed scales, this work provides preliminary benchmark distributions of motor dysfunction severity and frequency. Subsequent longitudinal follow-up studies will evaluate within-person trajectories, test-retest reliability, and responsiveness to clinically meaningful change. This will support their eventual use as quantitative outcome measures for motor function in therapeutic trials for NDDs.^20^

## Results

### Cohort Overview

We analyzed motor phenotypes in 102 individuals with a neurodevelopmental disorder secondary to a pathogenic or likely pathogenic variants in five NDD-associated genes and 31 unaffected sibling controls (controls) using unique, standardized motor assessments (Table 1). Among individuals with NDDs, *SCN2A* was the largest group (n = 48), followed by *KCNQ2* and *CSNK2A1* (n = 20 each), *CACNA1A* (n = 8), and *SCN8A* (n = 6) (Table 1). Participants were recruited from multidisciplinary clinics at UTHealth Houston or from gene-specific advocacy events across the United States and Europe between August 2024 and September 2025. The age at assessment ranged from five months to 38 years (median 10 years; interquartile range [IQR] 5 – 15 years). Two controls and seven affected individuals were under three years of age. The cohort was predominantly White (~78%), with limited representation from Asian, Hispanic, and Black participants, reflecting known recruitment biases in rare disease cohorts.^21^

**Table 1.** Demographics. Baseline demographic characteristics of this cohort. Units for each value are specified alongside the respective characteristic.

| Characteristic | Total |
|---|---|
| Recruitment Period | 8/2024 – 9/2025 |
| Total number of individuals – n | 133 |
| Genotype – n (%) |  |
| Control | 31 (23.3) |
| *CACNA1A* | 8 (6.1) |
| *CSNK2A1* | 20 (15.0) |
| *KCNQ2* | 20 (15.0) |
| *SCN2A* | 48 (36.1) |
| *SCN8A* | 6 (4.5) |
| Sex – n (%) |  |
| Female | 60 (45.1) |
| Male | 73 (54.9) |
| Age at time of assessment – n (%) |  |
| 0 – 2 \| Infancy | 9 (6.8) |
| 3 – 5 \| Early Childhood | 31 (23.3) |
| 6 – 10 \| Late Childhood | 32 (24.1) |
| 11 – 18 \| Adolescence | 35 (26.3) |
| >18 \| Adulthood | 22 (16.5) |
| N/A | 4 (3.0) |
| Age range – years (median; IQR) | 0.5 – 38 (10; 5 – 15) |
| Ethnicity – n (%) |  |
| Asian | 9 (6.8) |
| Black | 4 (3.0) |
| Hispanic | 16 (12.0) |
| White | 104 (78.2) |
| Wheelchair status among affected individuals – n (%) |  |
| Yes | 29 (28.4) |
| No | 73 (71.6) |

### Individuals with NDDs exhibit a broad range of motor impairments

Motor phenotypes were assessed using standardized clinical scales for spasticity (Modified Ashworth Scale [MAS]),^22^ dystonia (Burke-Fahn–Marsden Scale [BFMS]),^23^ and ataxia (Scale for the Assessment and Rating of Ataxia [SARA]^24,25^. We first screened for the presence of motor dysfunction in the combined cohort of individuals with NDDs and controls across five domains (Figure 1A, Table 2). Speech dysfunction, including dysarthria and language-related dysfunction, was most prominent, occurring in 90% of individuals with NDDs. Gross motor and coordination deficits, and dystonia were observed in 82.4%, 87% and 83.3% of individuals with NDD, respectively, whereas spasticity was less common (42.2%; Figure 1A). In contrast, controls exhibited minimal impairment (0 - 12.9% across domains; Figure 1A). Overall, these results demonstrate that the scales detect abnormal motor phenotypes, and that minimal dysfunction is seen in controls. Given this low frequency of motor dysfunction across domains in controls, subsequent analyses focused on assessments involving individuals with NDDs.

**Table 2.** Parameter Overview and Categorization. Categorization of all parameters from the SARA, Fahn-Marsden and Modified Ashworth scales into subdomains and domains based on body region or functional grouping for simplified phenotypic description.

| Scale | Parameter | Subdomain | Domain |
|---|---|---|---|
| SARA – Ataxia | Sitting | Gross Motor | Gross Motor |
|  | Stance |  |  |
|  | Gait |  |  |
|  | Finger Chase | Coordination | Coordination |
|  | Nose-Finger Test |  |  |
|  | Fast-Alternating Hand Movement |  |  |
|  | Heel-Shin Slide |  |  |
|  | Speech | Ataxic Dysarthria | Speech |
| Burke-Fahn-Marsden – Dystonia | Speech and Swallowing | Speech Dystonia |  |
|  | Eyes | Orobuccal Dystonia | Dystonia |
|  | Mouth |  |  |
|  | Neck | Axial Dystonia |  |
|  | Trunk |  |  |
|  | Upper Extremity (Right/Left) | Extremity Dystonia |  |
|  | Lower Extremity (Right/Left) |  |  |
| MAS – Spasticity | Neck Flexion | Axial Spasticity | Spasticity |
|  | Neck Extension |  |  |
|  | Elbow Flexion (Right/Left) | Upper Extremity Spasticity |  |
|  | Elbow Extension (Right/Left) |  |  |
|  | Hip Flexion (Right/Left) | Lower Extremity Spasticity |  |
|  | Hip Extension (Right/Left) |  |  |
|  | Knee Flexion (Right/Left) |  |  |
|  | Knee Extension (Right/Left) |  |  |

**Figure 1.**
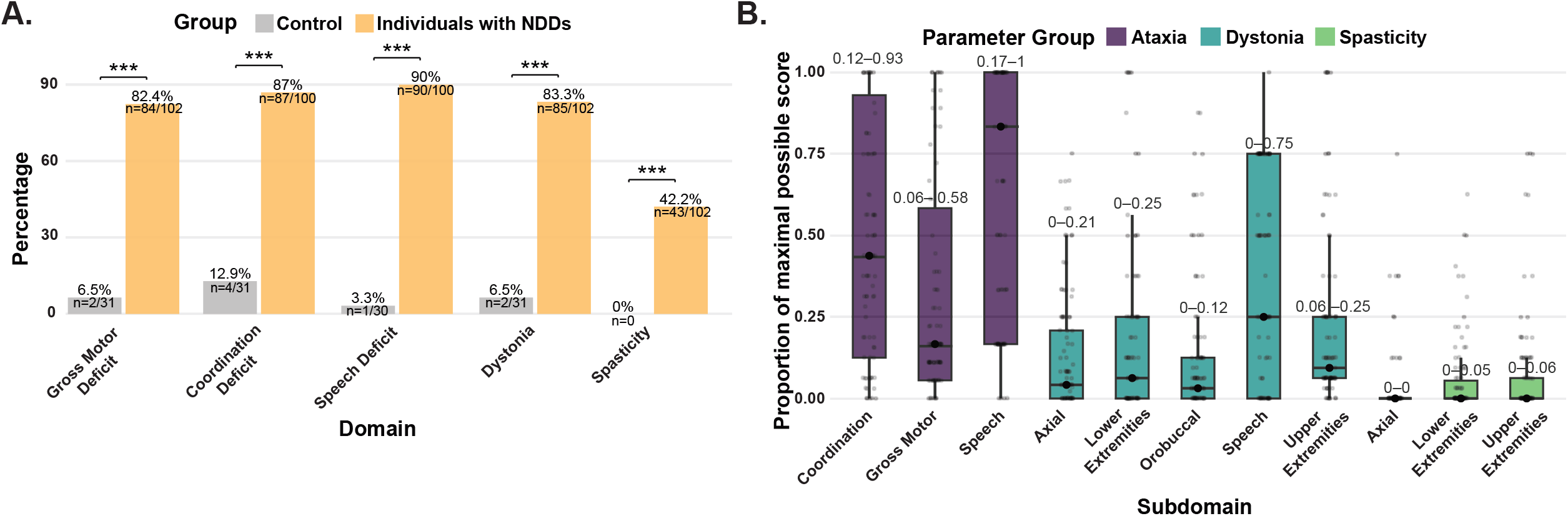
A. Frequency of deficits in motor domains across all assessments in individuals with NDDs (orange) and controls (grey). Deficits were determined as any assessment with a score ≥1 in any parameter within the domain (Table 2). Bars represent percentages with counts and each sample size excluding participants with missing data are shown within bars. Group comparisons were performed using Fisher’s exact test with BH correction: * <0.05, ** <0.01, *** <0.001, **** <0.0001. B. Boxplots depicting the proportion of maximal possible score attained for motor subdomains grouped into spasticity, dystonia, and ataxia. To obtain the proportion, sums of individual parameters of each domain were divided by the maximal possible score. Points represent individual patients, circles within the boxes indicate the medians, the edges of the boxes the Interquartile range (IQR); the IQR is also provided as a number above each box.

### Motor impairment severities follow subdomain-specific patterns

To capture heterogeneity of motor dysfunction in body part involvement, we assessed impairment across granular subdomains in all individuals with NDDs (Table 2; Figure 1B). Speech dysfunction as scored by the SARA which includes ataxic dysarthria and anarthria unatribbutable to a ataxia specifically, had the highest score distribution, with a median score of 0.83 (IQR 0.17 – 1), indicating maximal impairment in at least half of the cohort. Coordination deficits (median 0.44, IQR 0.12 – 0.9) and gross motor deficits had lower score distributions (median 0.17, IQR 0.06 – 0.58). Speech dysfunction as scored by the BFMDS was also prominent (median 0.25, IQR 0 – 0.75), whereas axial, limb, and orobuccal dystonia remained in the lower range (medians 0.03 – 0.09, IQR 0 – 0.25). Spasticity parameters had near floor-level distribution across axial and limb regions (Figure 1B). To asssessed consistency between raters, we evaluated inter-rater reliability for BFMS assessments. Agreement between the primary rater and an expert committee was excellent (ICC(2,1) = 0.90, 95% CI 0.84-0.94; n = 57 complete paired assessments; Table S2). We also estimated ceiling and floor effects for the three scales to identify measurement-related confounding. All spasticity-related subdomains demonstrated a pronounced floor effect (70–86%). In contrast, we observed minimal floor effects for ataxia (10-16%) and dystonia (22-55%) subdomains (Figure S1). Ceiling effects were generally low across subdomains, except for the speech parameter scored by the SARA (~50%) (Figure S1).

### Genotypes exhibit subdomain-specific motor impairments

We next explored gene-associated patterns across the cohort using group-level z-score averaging and descriptive statistics (Figure 2 and S2). No formal statistical comparisons were performed across genotype groups due to power constraints given the small gene-specific cohorts; confidence intervals are provided to illustrate variability in these exploratory estimates. Individuals with *SCN2A* variants demonstrated higher mean z-scores across most subdomains, suggesting greater overall severity relative to other genes in our analysis (Figure 2). Mean z-scores for ataxia coordination and speech (Atax_Coord: mean z = 0.28, 95% CI 0.03-0.54; Atax_Speech: mean z = 0.3, 95% CI 0.03-0.57; Table S3) indicated dysfunction in these domains with consistent deviation from the cohort mean. However, confidence intervals overlapped zero across several other domains, reflecting uncertainty in the magnitude of differences (Table S3). In contrast, individuals with *KCNQ2* and *CACNA1A* variants exhibited domain-specific patterns with considerable uncertainty. *KCNQ2* was associated with higher mean z-scores in speech dystonia (Dys_Speech: mean z = 0.27, 95% CI −0.22-0.78), whereas *CACNA1A*-associated dysfunction was largely confined to gross motor ataxia (mean z = 0.19, 95% CI −0.47-0.9; Table S3). Individuals with *CSNK2A1* and *SCN8A* variants showed lower relative severity across subdomains in this cohort, although confidence intervals for most domains overlapped zero, indicating substantial uncertainty in these estimates (Figure 2 and S2; Table S3). Given the overall low cohort sizes, all genotype-related findings should be considered hypothesis-generating, highlighting selective domain-level differences rather than uniform genotype effects.

**Figure 2.**
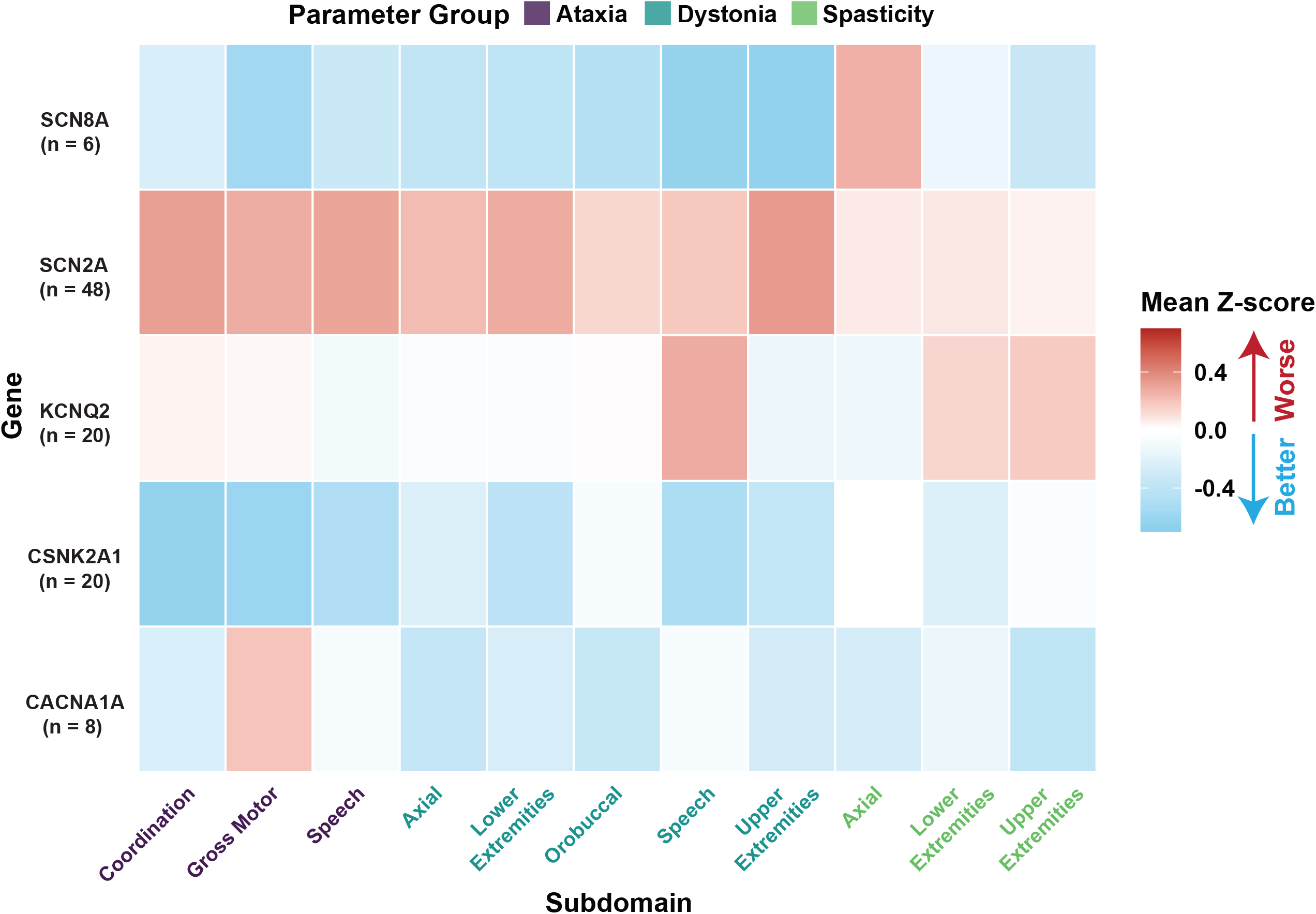
Heatmap depicting grouped mean z-scores across all parameter subdomains for individuals with variants in *CACNA1A, CSNK2A1, KCNQ2, SCN2A*, and *SCN8A*. Z-scores were standardized relative to the pooled affected cohort and averaged using each subdomain-assigned parameters followed by averaged at the gene level. Individual cells are color-coded by relative z-score, with red indicating higher z-scores (greater impairment) and blue indicating lower z-scores (less impairment). Sample sizes for each genotype group are indicated alongside gene labels. Cells represent descriptive estimates; no formal between-gene hypothesis testing was performed.

## Discussion

Motor dysfunction is widespread across monogenic NDDs but poorly defined and monitored.^26^ In this prospectively recruited, cross-sectional study, we quantify motor dysfunction frequency and severity in 102 individuals with monogenic NDDs using three complementary scales assessing ataxia, spasticity, and dystonia, with particularly deep quantification of individuals with *SCN2A, KCNQ2*, and *CSNK2A1*-related NDDs. Unlike prior studies utilizing retrospective chart review or video assessment, we utilize a standardized video protocol across five monogenic NDDs with domain-resolved quantification, providing greater phenotypic resolution and measurement rigor than categorical descriptions of motor dysfunction alone. Although gene-stratified results are exploratory, our observations suggest potential genotype-specific motor profiles that may inform the design of larger, adequately powered studies. Overall, our results provide crucial benchmark distributions of domain-specific motor dysfunction in monogenic NDDs. Subsequent longitudinal validation will support the development of motor endpoints for natural history studies and clinical trial readiness in rare diseases.

Prior studies have established the presence of motor dysfunction in individuals with monogenic NDDs. Van der Veen et al. reported the diversity of movement disorders in 77 individuals covering 38 genetic DEEs with known movement disorders, identifying both hypo- and hyperkinetic disorders using caregiver-submitted and archived clinical videos.^6^ Montanucci et al. similarly provide frequency estimates of motor phenotypes in 1210 probands across 87 NDD-associated genes without in-depth quantification.^27^ More recently, Bernardi et al summarized the growing body of knowledge suggesting motor dysfunction is prominent in epilepsy-associated NDDs.^28^ Gene-specific studies also suggest disease mechanism-dependent motor dysfunction but they primarily describe motor phenotypes as comorbidities rather than quantifying frequencies and genotype-associated expression patterns.^10,29–33^ A key retrospective study by Mastrangelo et al. provides important reference ages for motor phenotypes.^34^ Thus, the major gap is no longer whether motor dysfunction occurs, but how to measure its severity so that it can be used in natural history studies and trials. Our work targets this gap by applying validated, domain-resolved scales across a wide age, building a foundation for systematic motor outcome assessment in NDDs.

Our analyses demonstrate low frequency and lower score distributions of spasticity across the NDD cohort. Spasticity reflects velocity-dependent hypertonia arising from corticospinal tract injury, classically associated with perinatal brain injury.^35^ In monogenic NDDs, focal corticospinal injury is not typically expected or rare, in line with our observation of an overall low spasticity burden. Accordingly, the pronounced floor effect observed for the MAS may reflect a genuinely low prevalence of spasticity in this population. However, given the substantial floor effects across spasticity-related subdomains (70–86%), reduced sensitivity of the scale to detect milder forms of hypertonia cannot be excluded. These findings should therefore be interpreted cautiously, and spasticity-related results may be influenced by both true low frequency and measurement limitations.

Hypotonia is the most frequently reported motor phenotype across NDDs and developmental and epileptic encephalopathies.^9,36^ We did not systematically measure hypotonia due to the lack of standardized scales applicable across development, but it was clinically apparent in most assessments, and we hypothesize that our data provides indirect evidence for this observation. A large proportion of our cohort exhibited dystonia; multiple lines of evidence suggest that hypotonia may predispose to secondary dystonic posturing, particularly of distal extremities.^37^ In line with this hypothesis, dystonic features were often discrete and limited to distal extremities, possibly also reflecting abnormal sensory processing, which can contribute to focal dystonia.^38,39^ Together, these factors provide a biological basis for the higher severity of dystonic features and the possible association with widespread hypotonia observed in this cohort.

The use of the SARA across a broad developmental range introduces important limitations, as several components (e.g., gait and stance) were originally validated in ambulatory populations without developmental disorders.^24^ For non-ambulatory participants, maximum scores were assigned for mobility-dependent items, which may contribute to ceiling effects and reduce sensitivity to gradations of dysfunction. For pre-ambulatory participants, these items were not scored and designated as not applicable. However, in the absence of validated motor scales applicable across both ambulatory and non-ambulatory individuals, this approach allowed for pragmatic stratification of motor severity across the cohort.

Speech dysfunction was prominent across our cohort, including among individuals with milder limb or axial motor dysfunction. The SARA speech item assesses the intelligibility of spoken language as a measure of ataxic dysarthria. However, it has not been validated for individuals who have not acquired speech because of developmental impairment.^25^ In these participants, a high score cannot be attributed specifically to ataxic dysarthria and may instead reflect language or cognitive impairment.^40^ The pronounced ceiling effect in our cohort may partly reflect this limitation; the item also does not capture vocalizations, gestures, or other forms of communication particularly relevant to cognitively impaired individuals.^41–43^ We therefore interpreted these scores as measures of observed function, not isolated evidence of cerebellar impairment. Approximately one third of individuals in our cohort also showed elevated scores for speech on the BFMS, suggestive phonetic or vocalization deficits as well. However, it also cannot adequately distinguish whether speech impairment is related to cognitive or motor vocalization. Overall, the value of speech assessment as a functional outcome measure cannot be understated. Granular, developmentally appropriate tools to disentangle linguistic and cognitive determinants of speech impairment in NDDs are much needed future optimizations to our protocol.^44^

While limited by sample size, our data suggest gene-associated patterns of motor dysfunction. Individuals with *SCN2A* variants demonstrated higher mean z-scores across multiple subdomains. These findings align with the known phenotypic spectrum of *SCN2A*-related disorders, which range from severe developmental and epileptic encephalopathies (DEEs) to milder behavioral phenotypes.^29^ *KCNQ2* and *CACNA1A* variants exhibited domain-specific patterns with substantial uncertainty. *CSNK2A1*-related NDD exhibited comparatively milder global motor impairment, with language and cognitive features frequently predominating, findings directionally consistent with observed clinical patterns.^45^ Taken together, our results highlight selective domain-level differences rather than uniform genotype effects and should be interpreted as hypothesis-generating for larger gene-stratified studies.

Key drawbacks that must be mentioned are that our cohort may not be representative of all NDDs given its limitation to five genes and the pre-dominance of channelopathy-related genes. Moreover, the lack of variant-level stratification within the gene groups may also limit the generalizability of our findings. For instance, *SCN2A* variants exhibit substantial functional heterogeneity, with gain-of-function variants typically associated with severe DEE and loss-of-function variants more commonly linked to milder neurodevelopmental phenotypes.^46^ Pooling these mechanistically distinct variant classes within a single gene-level group may obscure meaningful differences in motor dysfunction expression and contribute to variability in the observed severity patterns. This may partially explain the variability and confidence intervals overlapping zero observed within the *SCN2A* group. Accordingly, the gene-level estimates presented here should be interpreted as aggregate patterns rather than mechanism-specific effects. Future studies incorporating more homogeneous cohorts or stratification by variant-level biology, severity, and age of onset will be necessary to disentangle gene-specific effects from underlying clinical heterogeneity.

Our study has several other limitations that should be considered when interpreting these findings. First, recruitment from gene-specific advocacy events and a tertiary academic center likely introduce ascertainment bias. Participation in these settings requires travel and caregiver availability, likely limiting inclusion of individuals with more severe disabilities. These individuals are also more likely to receive ancillary therapy support, limiting generalizability to individuals without access to these services. Moreover, increased mortality among individuals with severe NDDs may contribute to an overrepresentation of milder phenotypes in older age groups. Therefore, age-associated findings should be interpreted cautiously. Our recruitment sites likely also introduced socioeconomic biases, reflected in the predominantly White cohort which limits external validity to racial and ethnic minorities. This limitation is common in rare disease research and underscores the need for strategies to improve accessibility and inclusivity across socioeconomic backgrounds.^21^ Further collaboration with patient-advocacy groups to address this gap, for instance by promoting virtual, in-person assessments may improve generalizability and access to larger, representative cohorts. Although missing data were infrequent, incomplete assessments occurred primarily because of behavioral intolerance, fatigue, or time constraints and therefore may not have been completely random. However, fewer than 5% of observations were excluded for most analyses (Table S4), reducing the likelihood that missingness substantially influenced the overall findings. Previous studies have demonstrated the presence of additional motor phenotypes such as chorea and myoclonus in patients with NDDs.^6,27^ Although we frequently observed these features, they were not systematically analyzed. Many of these phenotypes lack standardized severity scales that are broadly applicable across developmental ages. Our focus on ataxia, dystonia, and spasticity was therefore driven by the availability of validated rating instruments, highlighting the need to develop scalable, quantitative tools to enable inclusion of a wider range of motor phenotypes as outcome measures. Additionally, motor assessments were scored by a single expert rater, supervised by developmental neurology and movement disorder specialists. Inter-rater reliability testing between the primary rater and an expert committee demonstrated excellent agreement for the BFMS (ICC(2,1) = 0.90). However, scales such as the MAS require in-person examination and cannot be reliably scored post-hoc using videos alone, limiting formal assessment of reliability for all scales. Future studies would benefit from incorporating protocols optimized for remote and asynchronous scoring to facilitate broader participation, and independent validation. Moreover, we did not systematically collect detailed longitudinal clinical data on potential confounders such as seizure burden, antiseizure medication exposure, or prior neurosurgical interventions. As many participants were recruited at advocacy conferences, comprehensive medical histories were not uniformly available, and we cannot rule out additional underlying structural brain abnormalities which may influence motor phenotypes and increase unmeasured heterogeneity within the cohort. Future studies incorporating structured clinical data collection and medication histories will be necessary to disentangle intrinsic genotype-associated motor dysfunction from treatment or comorbidity-related effects. Finally, we emphasize the limited sample size of our study, which may affect statistical power and generalizability; findings should therefore be interpreted within the context of an exploratory study of rare monogenic NDDs.

In summary, this study provides a much needed standardized, domain-resolved quantification of motor dysfunction across developmental stages in monogenic NDDs. Our findings demonstrate that motor assessment scales are straightforward to administer and can be reliably applied with appropriate training, making them well suited for routine clinical use and research. Their broader adoption as standard tools in clinical practice would substantially expand the availability of structured motor data across diverse settings. This framework establishes a foundation for longitudinal motor phenotyping and supports the development of quantitative motor outcome measures for natural history studies and emerging clinical trials.^47^

## Methods

### Cohort Recruitment

This prospectively recruited, cross-sectional observational study was approved by the Committee for Protection of Human Subjects of UTHealth Houston (IRB# HSC-MS-24-0370). All individuals with variants in NDD-associated genes and controls were prospectively recruited over 13 months (08/2024 – 09/2025) at the *SCN2A* and *CACNA1A* multidisciplinary clinics at UTHealth Houston and at six patient advocacy group conferences: FamilieSCN2A Foundation 2024 and 2025 (https://www.scn2a.org/), The Cute Syndrome Foundation 2024 (https://www.thecutesyndrome.com/), CSNK2A1 Foundation 2025 (https://www.csnk2a1foundation.org/), SCN2A/SCN8A Alliance 2025 (https://scn8a.eu/) and the KCNQ2 Cure Alliance 2025 (https://www.kcnq2cure.org/).

The inclusion criterion for individuals with NDDs was the presence of a pathogenic or likely pathogenic variant in the five NDD-associated genes we studied as per American College of Medical Genetics (ACMG) guidelines^48^. Seizure history was not required for inclusion and was not used to define the analytic cohort. Individuals with acute illnesses that could interfere with safe participation in the neurological examination were excluded from the study. Unaffected siblings of enrolled individuals with NDDs were included as controls without prior phenotype matching to facilitate pragmatic recruitment and reduce selection bias. Gene selection was driven by feasibility of recruitment, including patients evaluated in multidisciplinary clinics and at gene-specific advocacy events during the study period. As such, the cohort represents a pragmatic sample of individuals with monogenic NDDs rather than a biologically uniform group selected for mechanistic similarity.

The only inclusion criterion for controls was no known history of any NDD-associated variants. However, no explicit genetic testing was performed to rule out any incidental genetic findings. A formal *a priori* sample size calculation was not performed due to the rarity and heterogeneity of the studied NDDs. Recruitment was based on feasibility, and all eligible individuals during the recruitment period were invited to participate; therefore, we emphasize effect sizes and confidence intervals rather than post-hoc power estimates. All participants and/or legal guardians underwent a written informed consent process prior to study procedures. Age groups were defined as infancy (0-2 years), early (3-5 years) and late childhood (6-10 years), adolescence (11-18 years) and adulthood (>18 years) (Table 1).

### Data Collection and Curation

All consents, identifiable participant information and clinical data were stored in an instance of UTHealth RedCap to ensure strict adherence to data protection regulations. Each participant was assigned a unique identifier (UID); demographic and clinical data were de-identified prior to extraction from RedCap for analysis. Motor exam scores were collected in a blinded dataset containing the UID as the only identifier. The primary in-person rater was not blinded to recruitment context in all cases, whereas post-hoc expert committee BFMS re-rating was performed blinded to participant identity, genotype, and clinical information.

### Variant Pathogenicity Classification

Variant pathogenicity (re)classification was performed by variant analysis experts and genetic counselors in the Lal Group following ACMG criteria and using established *in silico* tools. We followed the recent variant curation expert panel (VCEP) guidelines developed for epilepsy-associated voltage-gated sodium channels (https://clinicalgenome.org/docs/clingen-variant-curation-expert-panel-vcep-protocol/).

### Motor Examination Setup

Assessments took place at UTHealth’s UT Physicians Clinic at the Texas Medical Center or in dedicated examination rooms at conference venues. The rater conducted examinations and team members assisted with documentation. We utilized two uniquely positioned Sony HDRCX405 Handycam Camcorders to record all standardized assessments for retrospective curation and independent rating for reliability testing. Caregivers were present throughout to ensure participants’ safety and comfort.

### Movement Rating Scales

Motor phenotypes were quantified using the Scale for the Assessment and Rating of Ataxia (SARA), the Burke-Fahn-Marsden Dystonia Rating Scale (BFMS), and the Modified Ashworth Scale (MAS) for spasticity. Domains and subdomains were established by stratifying individual parameters into clinically meaningful groups based on body region or motor function (Table 2). For non-ambulatory participants, SARA items requiring independent mobility (e.g., gait and stance) were assigned the maximum score, reflecting inability to perform the task. For pre-ambulatory participants, these items were not scored and were designated as not applicable. This approach was selected to preserve the ordinal structure of the scale and enable consistent severity categorization across the cohort in the absence of validated measures spanning the full developmental and functional spectrum. For the BFMS, final scores were calculated by multiplying raw scores with the provocation factor – defined as the presence of a phenotype at rest or only in certain ranges of motion – as described previously.^23^

### Handling of Missing Data

No imputation of missing data was performed. Analyses were conducted using an available-case approach, with observations included when the variables required for a given analysis were available to maximize sample size in this observational study. Consequently, denominators varied across analyses depending on the completeness of the relevant motor assessment.

Composite motor subdomain scores were derived by summing the available component scores within each subdomain, with available-item maximum used for proportion calculations. Participants for whom all component parameters within a subdomain were unavailable were assigned a missing value for that composite subdomain. When at least one component parameter was available, the composite score was calculated using the available observations without imputation.

Binary motor phenotypes were classified as present when at least one contributing parameter had a score ≥1 and absent when one or more contributing parameters were observed and all observed parameters had scores of 0. Binary phenotypes were considered missing only when all contributing parameters were unavailable. Accordingly, frequency estimates were calculated using the number of participants with available data for each phenotype as the denominator.

For exploratory gene-level analyses, patient-referenced z-scores were calculated only for observations with available motor scores and parameters demonstrating non-zero variability across the affected cohort. Parameters without measurable variability in the affected cohort were excluded from z-score standardization (Table S4). Inter-rater reliability analyses used only jointly scored items from both raters, and the primary ICC analysis was restricted to assessments with complete paired ratings across all nine BFMS items. No imputation was performed for missing paired ratings.

The number and proportion of observations excluded because of missing data at each stage of the analysis are summarized in Table S4.

### Motor Examination Scoring and Inter-Rater Reliability Testing

All motor assessments were scored by a single primary rater (S.B.) with formal training in the use of the SARA, BFMS, and MAS. Training consisted of supervised scoring of representative videos, scoring guideline review prior to study initiation under direct supervision by two pediatric neurologists (N.T. and G.V.A.) and a physical therapist (S.E.M.) specialized in pediatric movement disorders. This supervision occurred as part of routine multidisciplinary clinical care. Participants with incomplete motor examinations due to fatigue, behavioral intolerance, or time constraints were retained in the analysis. No participants were excluded post-enrollment; individuals unable to participate in motor assessments were not enrolled in the study. Post-hoc blind, independent re-rating of 59 BFMS assessments was conducted by a movement disorder expert committee (N.T., S.E.M. and R.F). Inter-rater reliability was evaluated using a two-way random-effects, absolute-agreement, single-measure intraclass correlation coefficient [ICC(2,1)]. Total BFMS scores were calculated using items scored by both raters. The primary analysis was restricted to assessments with complete paired ratings across all nine BFMS items; two assessments with one missing paired item were excluded. No imputation was performed. An available-case sensitivity analysis including all 59 assessments produced nearly identical results.

### Statistical Analysis

Statistical analyses were performed in R (v4.4.1) (RStudio v2024.0.2). For frequency analysis (Figure 1A), participants were classified according to the presence or absence of five broad motor phenotypes (gross motor deficit, coordination deficit, speech deficit, dystonia, and spasticity). Sums and percentages were calculated separately for controls and individuals with NDDs. Differences between groups were evaluated Fisher’s exact tests. P values were adjusted for multiple comparisons using the Benjamini–Hochberg (BH) procedure.

Motor subdomain severity in individuals with NDDs was summarized by calculating composite subdomain scores and expressing each as a proportion of the maximum possible score (Figure 1B). Continuous variables were summarized using medians and interquartile ranges (IQRs). Floor and ceiling effects for each subdomain (Figure S1) were defined as scores <5% or >95% of the maximum possible score, respectively.

To characterize gene-level motor phenotype patterns (Figure 2), controls were excluded and individual motor parameter scores were standardized relative to the pooled affected cohort. Parameter-level z-scores were averaged within assessments across parameters belonging to the same motor subdomain, followed by aggregation within genotype groups. Mean z-scores therefore represent the average deviation from the affected-cohort mean. Uncertainty was estimated using nonparametric bootstrap resampling (2,000 iterations) with percentile-based 95% confidence intervals (Table S3). Gene-level analyses were descriptive and exploratory; no formal statistical comparisons were performed because of limited and unequal genotype-specific sample sizes.

## Declarations

### Author Contributions

S.B., T.B. and D.L. conceived the project and designed analyses. S.B. conducted examinations, supported recruitment, performed all analyses and wrote the manuscript. T.B. performed variant analysis, assisted in examinations, and revised the manuscript. M.F., M.O-U., C.S., A.M., A.C., E.M., A.S., L.M., C.L., and C.H. led patient recruitment and assisted in examinations. C.L., and L.M. performed variant interpretation and modeling. G.T. established the database and assisted with documentation queries. L.S.M., S.E., B.B., S.S., K.C., L.M., S.J., P.F., J.S., and G.R. assisted with study design, patient recruitment, and logistical organization of assessments. S.E.M., N.T., and G.V.A. provided direct supervision and training of S.B. in the motor examination. S.E.M., N.T. and R.F. performed post-hoc video scoring for inter-rater reliability testing. V.K., W.F., J.L., M.S.P., S.M.W., S.E.M., S.A.V., S.D.L., G.V.A., I.E.S., N.T. and D.L. provided supervision revised the manuscript.

## Acknowledgments

Firstly, we acknowledge all families and affected individuals for participating in this study and supporting our work. We also thank all Lal lab members for their continued support and avid discussions. We thank other members of the Texas Institute of Restorative Neurotechnologies (TIRN) for suggestions and guidance. We appreciate Colleen Taylor for leading the IRB approval process and continued administrative support. We thank Karol Volante for assisting with IT-related affairs.

## Ethical Approval Statement

This study was approved by the Committee for the Protection of Human Subjects (CPHS), the Institutional Review Board of the University of Texas Health Science Center at Houston under #HSC-MS-24-0370. All patients underwent a formal informed consent process as per CPHS guidelines. The entire study complies by the CPHS policies which can be found here: https://www.uth.edu/cphs/policies/.

## Data Availability Statement

All de-identified participant data and the code for data analysis can be openly accessed on Github: https://github.com/Sunanjay/Motor_Assessment/

## Supplementary Data

**Figure S1.**
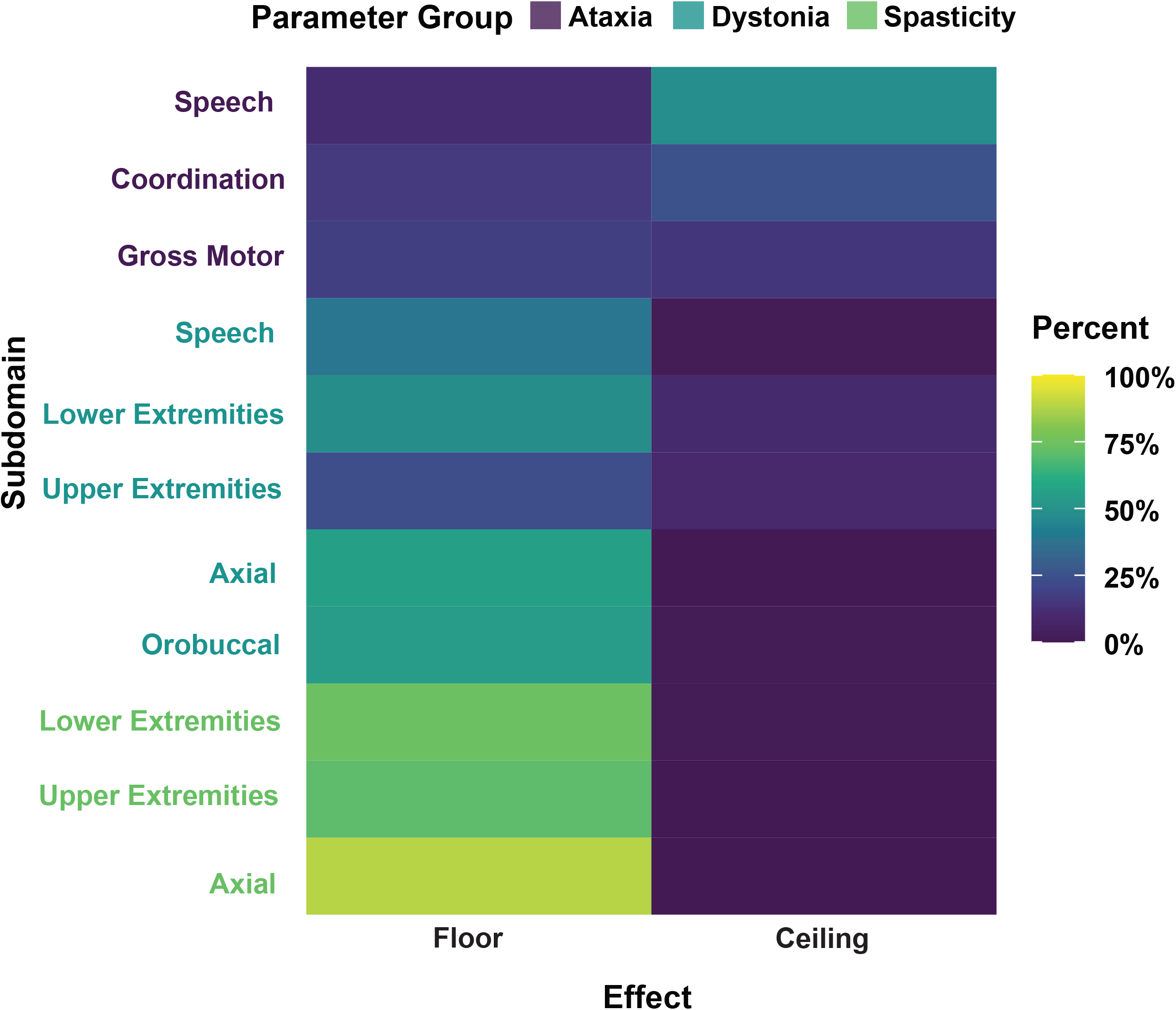
Heatmap showing the proportion of individuals demonstrating floor and ceiling effects across motor subdomains and parameter groups (ataxia, dystonia, spasticity). Floor and ceiling effects were defined as scores <5% and >95% of the maximum possible score, respectively. Rows represent individual subdomains, and columns denote floor and ceiling effects. Color intensity reflects the proportion of individuals, with higher values indicating a greater proportion of subjects at the floor or ceiling.

**Figure S2.**
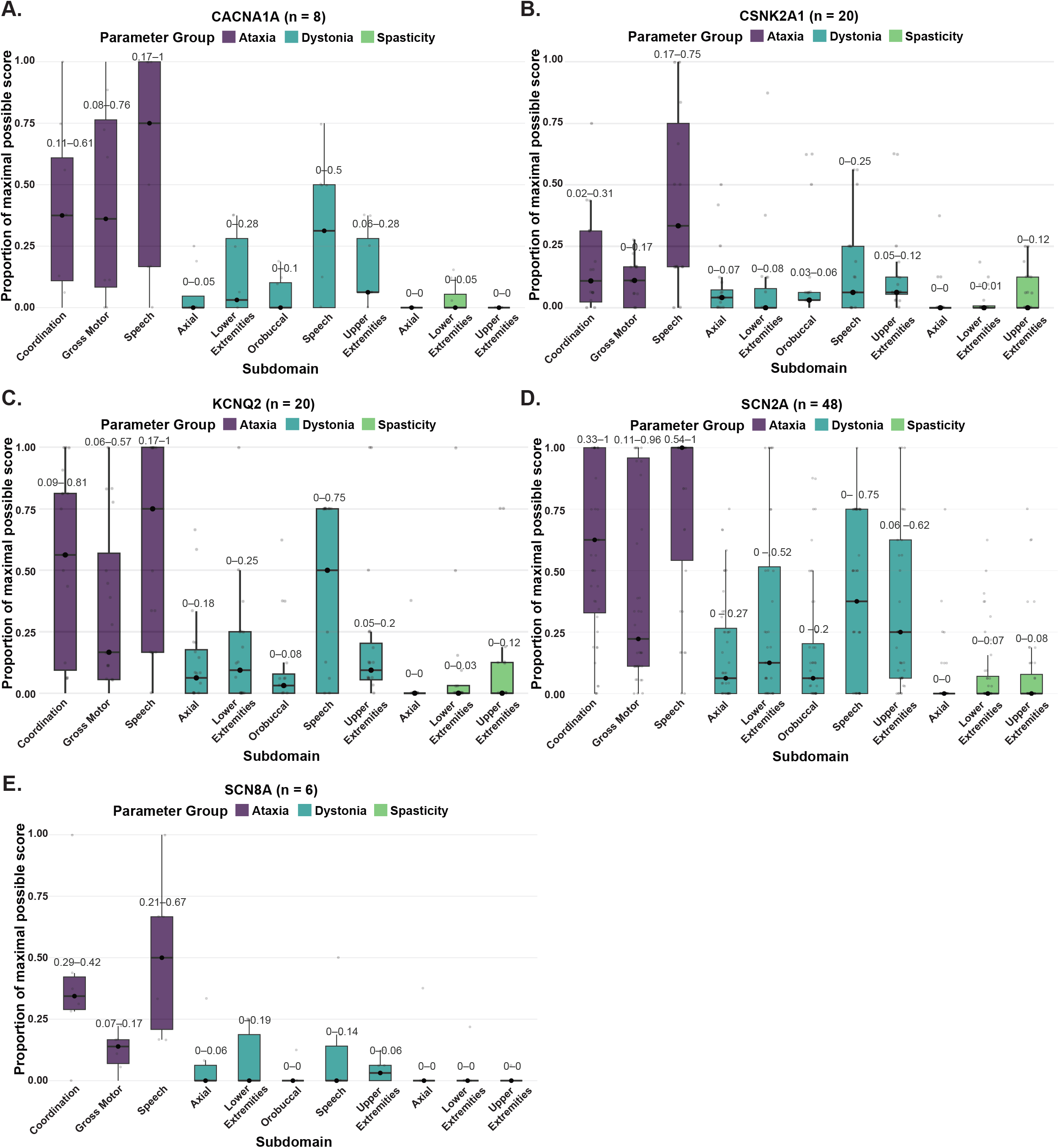
Boxplots illustrating the proportion of maximal possible scores attained for motor subdomains for individuals with variants in *CACNA1A* (A), *CSNK2A1* (B), *KCNQ2* (C), *SCN2A* (D), and *SCN8A* (E). Points represent individual patients, circles within the boxes indicate the medians, the edges of the boxes the Interquartile range (IQR); the IQR is also provided as a number above each box.

**Table S1.** Cohort sample size across age groups. Summary of cohort sample sizes for each gene in the respective age groups from infancy to adulthood as defined in Table 1.

|  | 0-2 | 3-5 | 6-10 | 11-18 | >18 | NA |
|---|---|---|---|---|---|---|
| Control | 2 | 5 | 2 | 14 | 5 | 3 |
| *CACNA1A* | 0 | 1 | 4 | 2 | 1 |  |
| *CSNK2A1* | 1 | 3 | 4 | 6 | 6 |  |
| *KCNQ2* | 1 | 5 | 8 | 3 | 3 |  |
| *SCN2A* | 5 | 15 | 11 | 11 | 5 | 1 |
| *SCN8A* | 0 | 1 | 1 | 2 | 2 |  |
| **Total** | 9 | 31 | 32 | 35 | 22 |  |

**Table S2.** Intraclass Coefficient Results. Interclass coefficient analysis statistics for inter-rater reliability comparison of the Fahn-Marsden Scale scoring. ICC(2,1) was selected as it estimates absolute agreement under a two-way random-effects model

| **Analysis** | **n** | **ICC(2,1)** | **95% CI** |
|---|---|---|---|
| Complete paired assessments | 57 | 0.904 | 0.84-0.94 |
| All available paired assessments | 59 | 0.903 | 0.83-0.94 |

**Table S3** Summary of mean z-scores, and confidence intervals for the exploratory genotype-level comparative analysis. Confidence levels were estimated by nonparametric bootstrap resampling with 2,000 iterations.

**Table S4** Summary of number of eligible observations, number analyzed, and number and percentage excluded because of missing data. Data is provided for each motor parameter, composite subdomain, binary phenotype, affected-cohort-standardized z-score, and inter-rater reliability analysis.

